# Mathematical estimation of COVID-19 prevalence in Latin America

**DOI:** 10.1101/2020.06.09.20126326

**Authors:** Marcos Matabuena, Pablo Rodríguez-Mier, Carlos Meijide-García, Víctor Leborán, Francisco Gude

## Abstract

After the rapid spread with severe consequences in Europe and China, the SARS-CoV-2 virus is now manifesting itself in more vulnerable countries, including those in Latin America. In order to guide political decision-making via epidemiological criteria, it is crucial to assess the real impact of the epidemic. However, the use of large-scale population testing is unrealistic or not feasible in some countries. Based on a newly developed mathematical model, we estimated the seroprevalence of SARS-CoV-2 in Latin American countries. The results show that the virus spreads unevenly across countries. For example, Ecuador and Brazil are the most affected countries, with approximately 3% of the infected population. Currently, the number of new infections is increasing in all countries examined, with the exception of some Caribbean countries as Cuba. Moreover, in these countries, the peak of newly infected patients has not yet been reached.

## 1 Main

In late December 2019, Chinese scientists isolated a novel coronavirus that was named as severe acute respiratory syndrome coronavirus 2 (SARS-CoV-2), and the relevant disease was called coronavirus-2019 disease (Covid-19) [1]. Since then, the dramatic increase in cases has posed numerous challenges to even the most sophisticated and advanced health systems, which led the World Health Organization (WHO) to declare the outbreak a pandemic in March 2020 [2]. Two months later, more than 2.4 million cases and over 143, 000 deaths have made the Americas the epicenter of the COVID-19 pandemic.

After North America, the sub-region with the highest number of confirmed cases and deaths reported to date remains South America. As of 28 May, the 10 countries in this sub-region have reported a total of 721, 893 confirmed cases, including 35, 412 deaths, which represents 30% of the total cases and 25% of the total deaths in the region of the Americas [3].

In the last decades, almost all of the South American countries introduced health financing and organizational reforms to strengthen health systems and to progress to universal health coverage. Despite improvements, inequities persist, and South America is the most inequitable region in the world, where there are disparities in social determinants of health [4]. The WHO risk assessment for Covid-19 is considered very high, mainly because the community transmission is ongoing in these territories; there is limited testing capacity; medical supply shortages; stringent population-level control measures are unlikely to be sustainable in the long-term; uncertainty about the resurgence of outbreaks following the relaxation of stringent control measures; the impact due to interruption of routine health services, routine vaccination and other disease control programs such as measles, polio, tuberculosis, and malaria, etc.

Given the ongoing reporting of Covid-19 cases in countries and territories in the Region of the Americas, PAHO/WHO continues to reiterate and update recommendations to support all Member States on the measure to manage and protect against the disease. A recent document developed by WHO to provide a practical guide to be used by national authorities states that Covid-19 surveillance data are essential to calibrate appropriate and proportional public health measures [5]. However, as we have pointed out, in Latin America, large-scale testing to quantify the spread of Coronavirus is an unrealistic methodology.

As an alternative to overcome this challenge, we can see two different strategies in the literature: i) survey data with geolocation technology as proposed by the team led by Eran Segal in Israel [6], ii) mathematical models to estimate the spread of the epidemic [7, 8]. The first option is not realistic in our context because mobile devices are not as widespread as in other places. As for the second point, many mathematical models were proposed in the literature, but based on our previous observation; we should consider using another source of information to fit the models instead of infection records [9].

In a recent paper [10], we proposed a new framework to estimate the prevalence of the virus in the population-based on mortality records. Our proposal was applied successfully in many countries and cities. After the first large-scale seroprevalence studies in different cities and countries, we confirm the accuracy of the previous initial estimations about percent of infections in each localization (more information available in https://covid19-modeling.github.io/). We refer to the original reference for technical mathematical details about the models.

This paper aims to show our estimate of seroprevalence as of May 27 in Latin America. We show only the complete results in Brazil and Peru due to space restriction, with the average infection estimate for most Latin American countries on a map. A more detailed description of the results can be found in the Supplementary Material and on the website related to the project (https://covid19-modeling.github.io/).

Before start, we introduce the main variables estimated by our model.

- *I*_1_(*t*): Number of infected individuals who are incubating the virus on day *t*.
- *I*_2_(*t*): Number of infected people who have passed the incubation period and who: i) does not show symptoms or ii) symptoms are mild on the day *t*.
- *I*_3_(*t*): Number of infected people who have passed the incubation period and do show severe symptoms on day *t*.
- *R*_1_(*t*): Number of recovered cases still able to infect day *t*.
- *λ*(*t*): Number of new infection on day *t*.

In addition, to increase the interpretability of the results, we show our adjusted model of death records and the total number of infections in each country under consideration.

Results in Brasil and Peru are shown in Figures 1 and 2. Averaged estimations of seroprevalence in other Latin American countries are shown in Figure 3.

**Figure 1:**
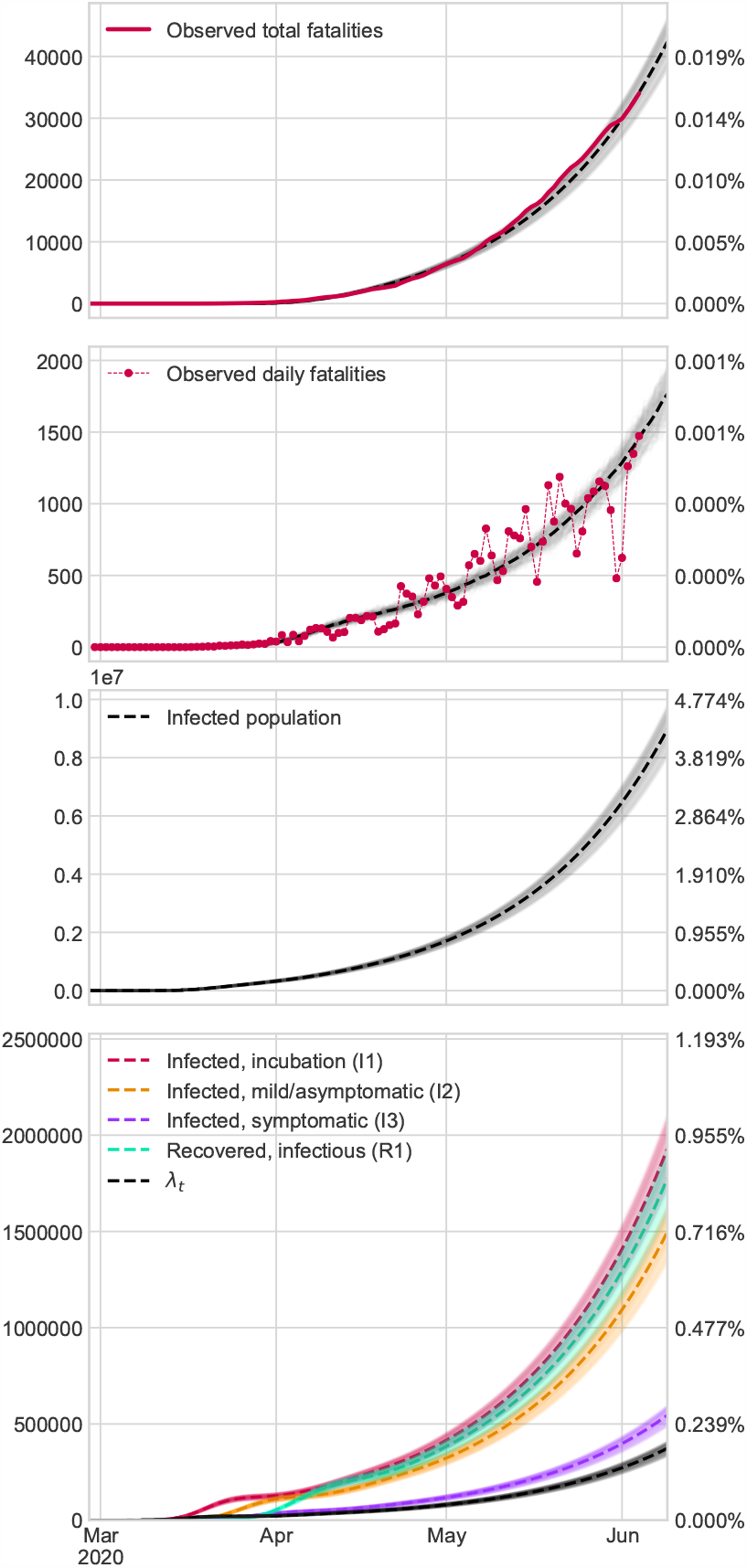
Brazil results by our mathematical model. We assume that there are as many fatalities as in the official records.

**Figure 2:**
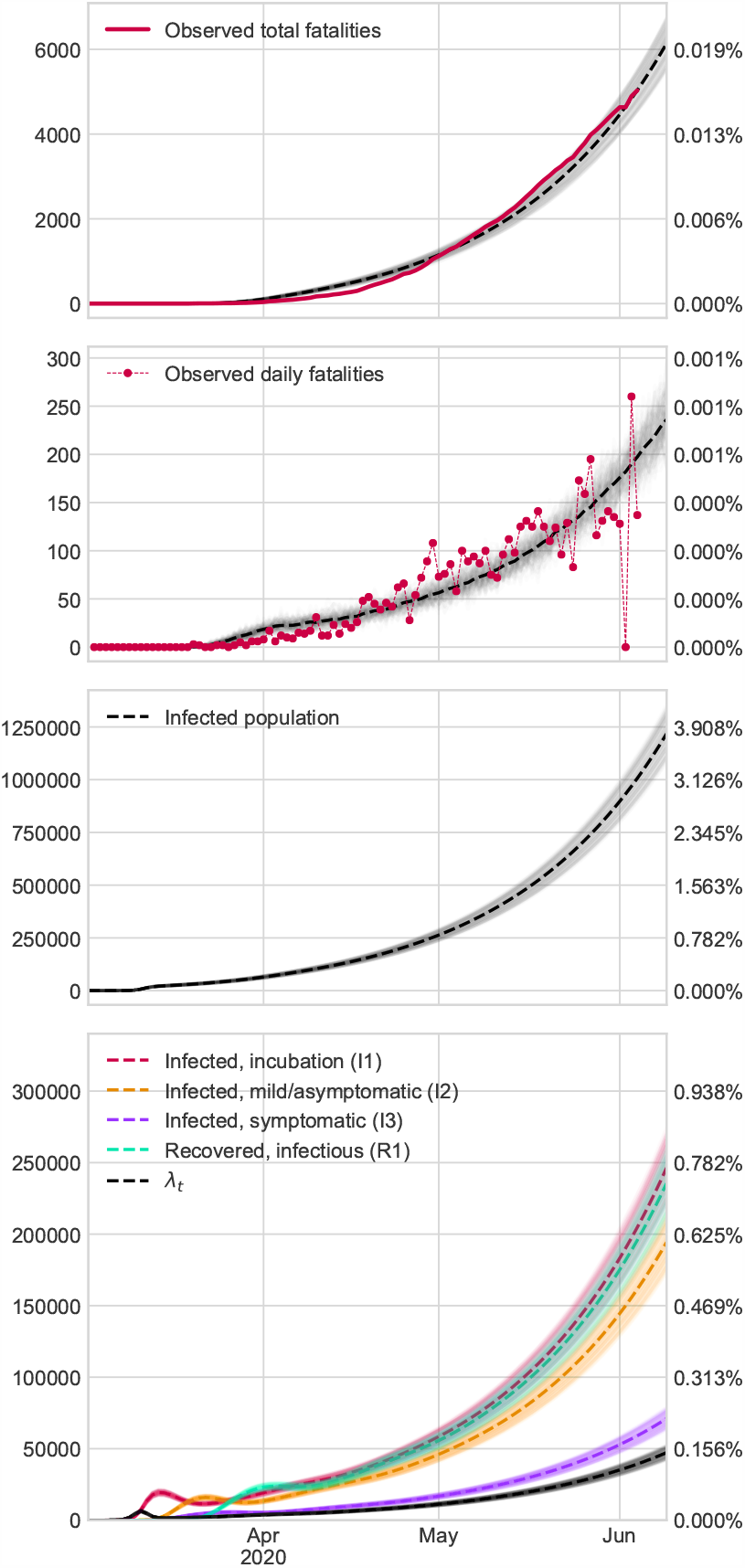
Perú results by our mathematical model. We assume that there are as many fatalities as in the official records.

**Figure 3:**
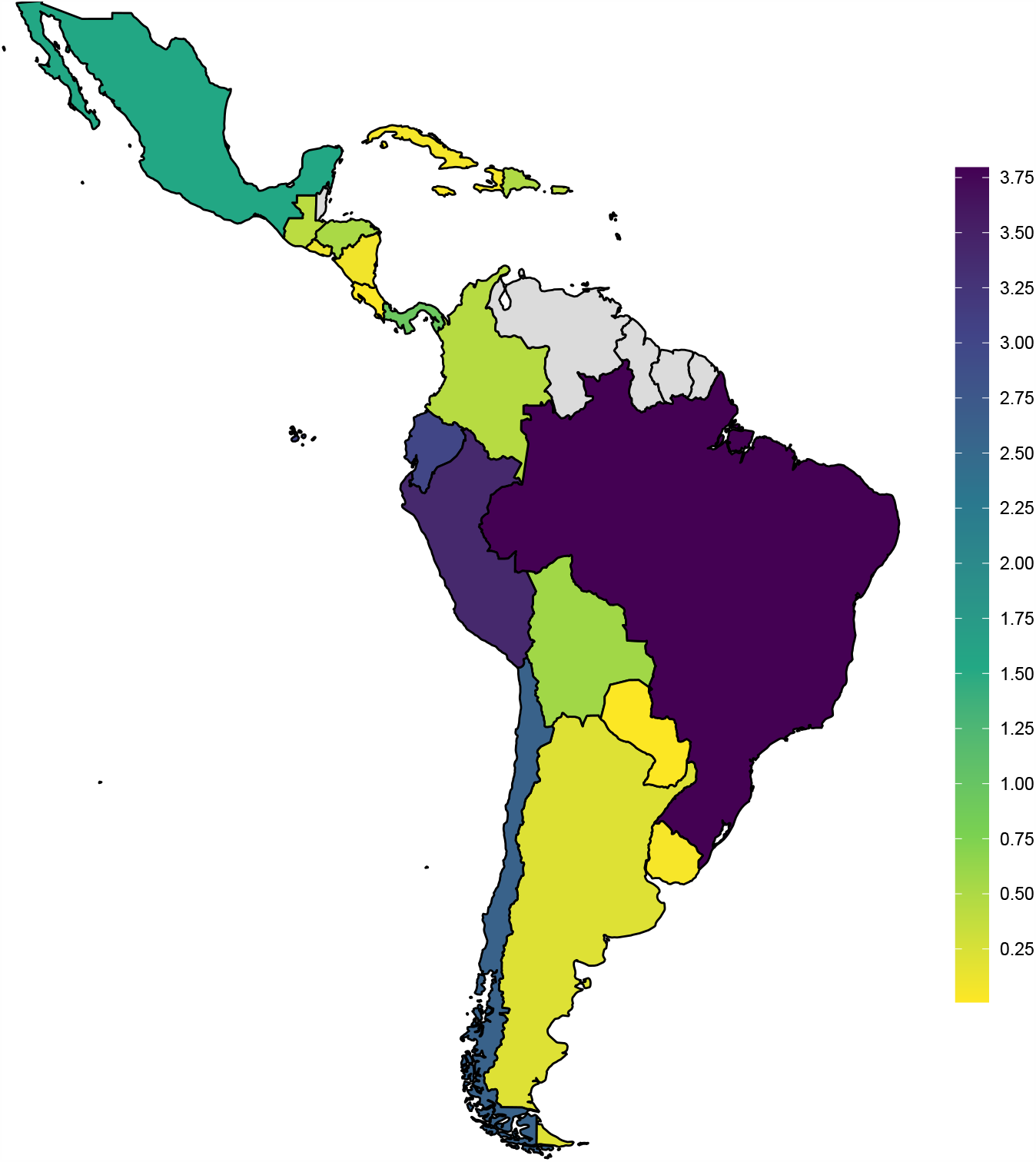
Map representing average estimations of seroprevalence percentage across several countries in Latin America. Gray colours indicates bad data quality or not enough data to fit models(colorblind-friendly palette)

As can be seen, there are two different groups regarding epidemic evolution. The first group corresponds to Central American and Caribbean countries such as Cuba and Puerto Rico, where the situation seems to be under control. The rest of the countries are in the other group where the rate of new infections is growing, and this may indicate that an epidemic situation is at its worst. In particular, countries with most infections are Ecuador, Brazil, Chile, and Peru, with an incidence of between 2.5 and 4% of the population. Interestingly, in Argentina and Colombia, the estimated number of infections is low, despite the geographical previous to the above countries.

The number of infections estimated by the model in Latin America is lower than in European epidemiological studies. However, in order to rigorously compare the spread of epidemics, we should perform the analyses under the same conditions. In Latin America, the number of new cases is increasing, and blocking measures or social distance cannot be so strict in Latin America or difficult to implement in some settings, for example, in the areas of highest poverty. It is therefore foreseeable that in the immediate future, the number of infections maybe even higher.

One possible limitation of our estimates is that the number of deaths may be higher than the government data reflect. A precise correction requires the use of previous mortality records, but this information is not always available in most of the countries under consideration. In fact, we excluded Venezuela and other countries in the analysis due to the poor quality of the data. In this context, we should be more cautious with estimates in Latin American countries than in European countries. In our estimations, we assumed an infection mortality rate (IFR) of 1.2%. Recent research [11] establishes that the IFR can range from 1 *−*1.4%; however, we must analyze the above factors on the demographic structure, epidemiological profiles, and quality of the health system. Further refinement is not possible due to unreliable data. From a public health perspective, our model helps to understand deeper insights as the virus spreads in these countries and to obtain important information such as the estimated seroprevalence of the population in a simple, low-cost procedure, which can be helpful to guide the next actions [7]. We are currently making efforts to maintain up-to-date estimates in the different countries of Latin America and other parts of the world on our website (https://covid19-modeling.github.io/). Figure 4 shows a screenshot with the information offered on the web for the particular case of Puerto Rico.

**Figure 4:**
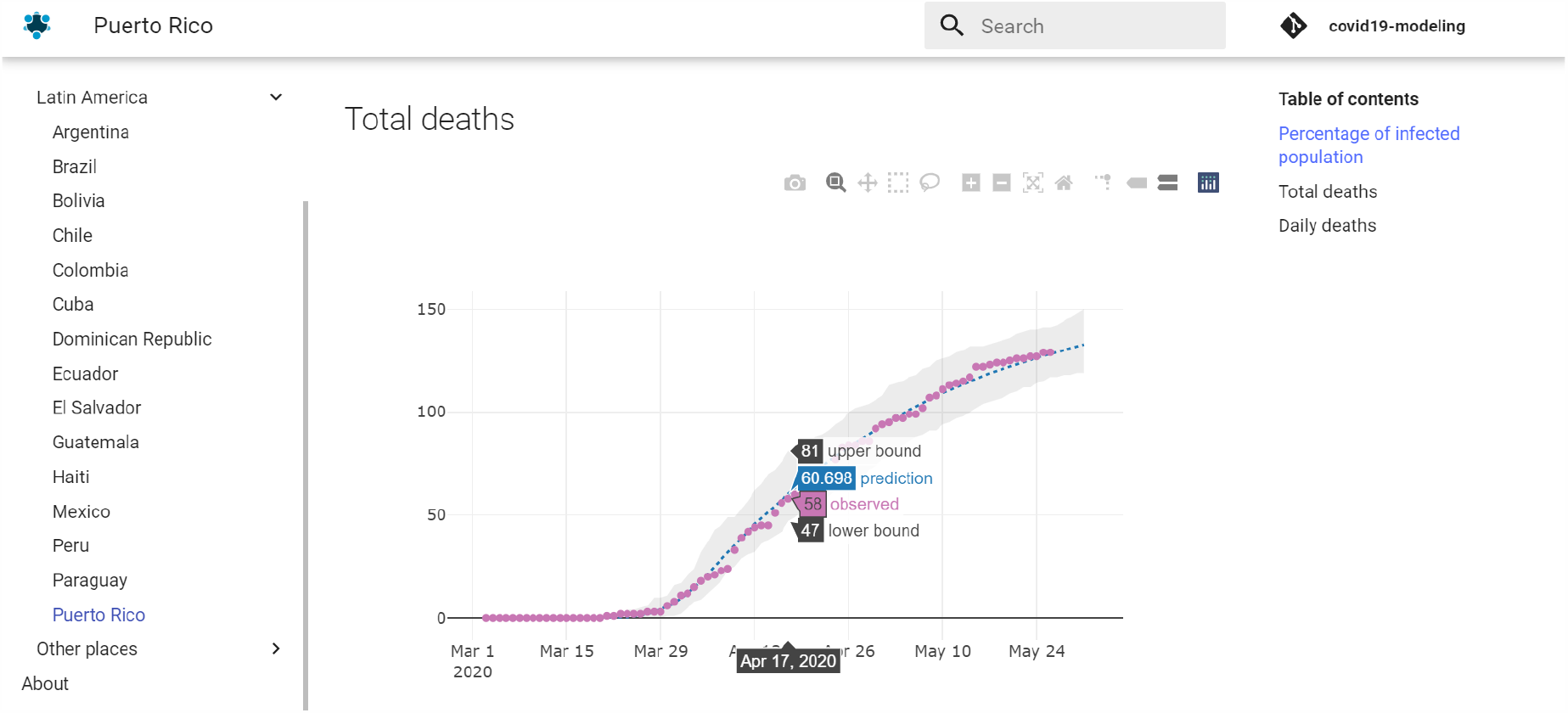
Screenshot of our dynamically updated webpage containing the results of worldwide estimations

**Figure 5:**
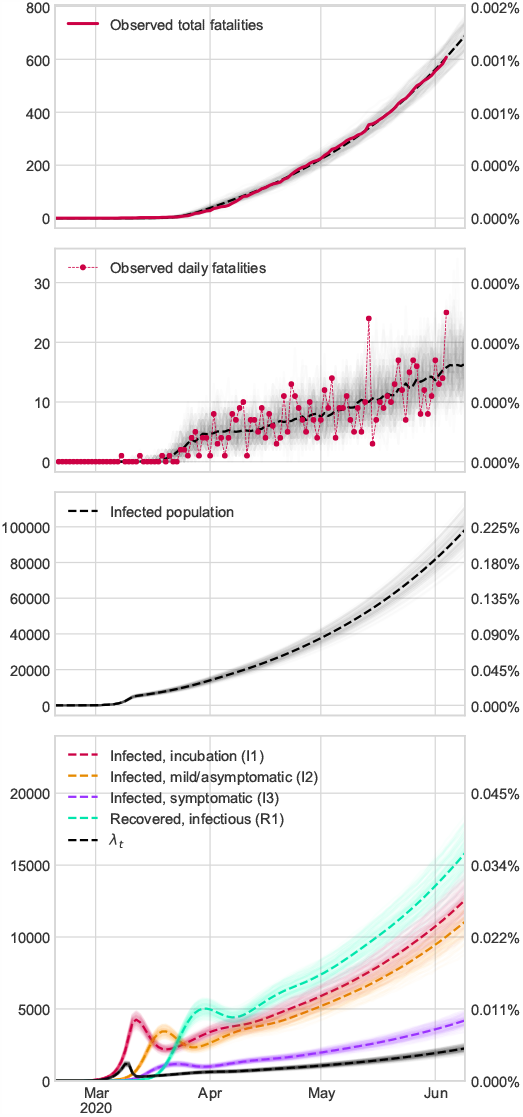
Results in Argentina: we assume that there are as many fatalities as in the official records.

**Figure 6:**
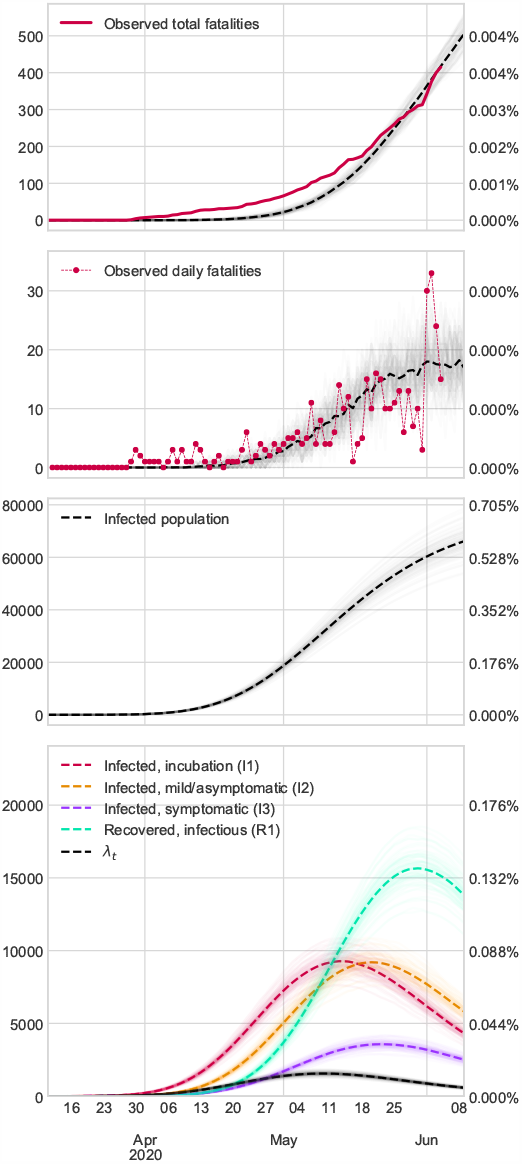
Results in Bolivia: we assume that there are as many fatalities as in the official records.

**Figure 7:**
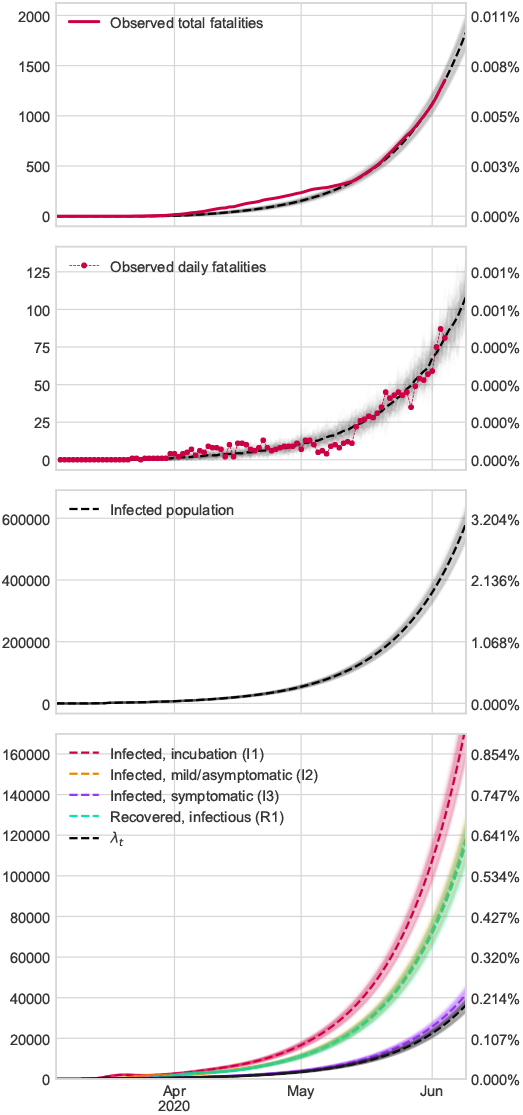
Results in Chile: we assume that there are as many fatalities as in the official records.

**Figure 8:**
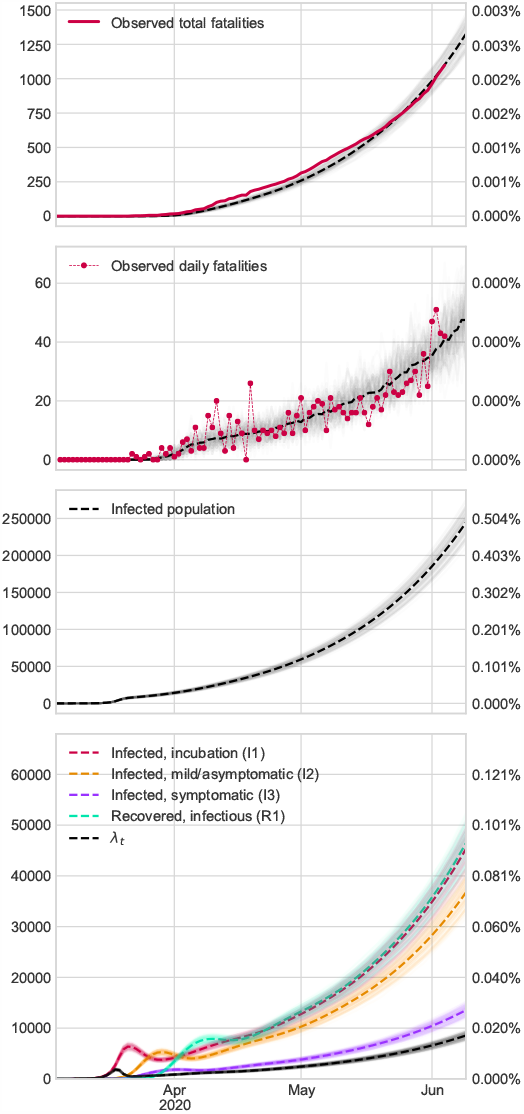
Results in Colombia: we assume that there are as many fatalities as in the official records.

**Figure 9:**
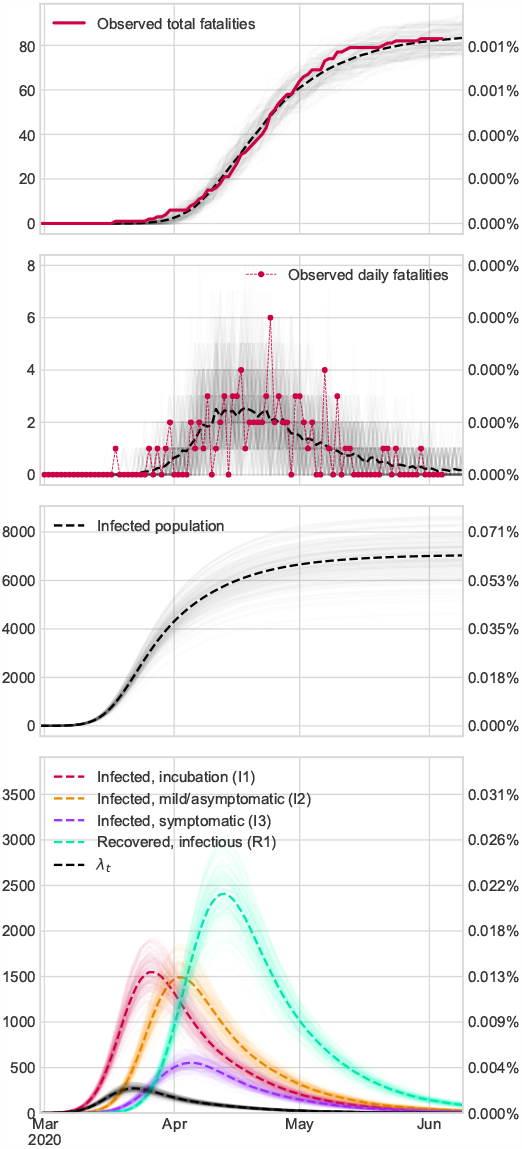
Results in Cuba: we assume that there are as many fatalities as in the official records.

**Figure 10:**
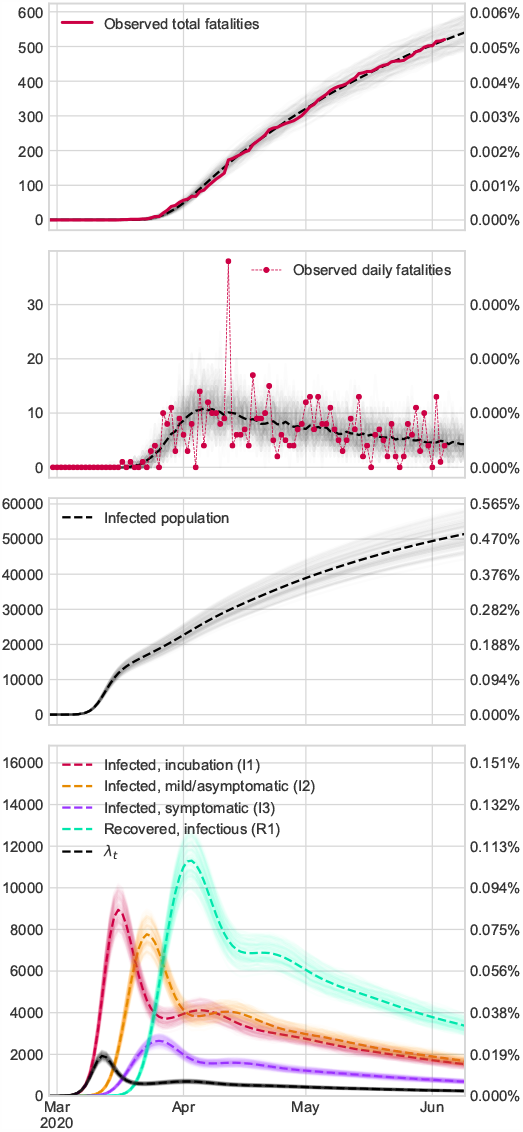
Results in República Dominicana: we assume that there are as many fatalities as in the official records.

**Figure 11:**
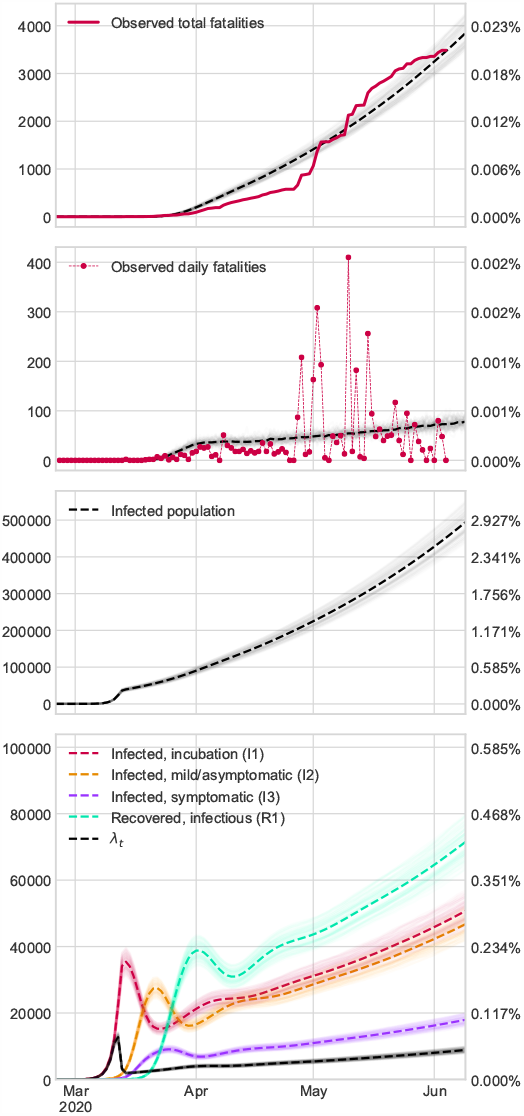
Results in Ecuador: we assume that there are as many fatalities as in the official records.

**Figure 12:**
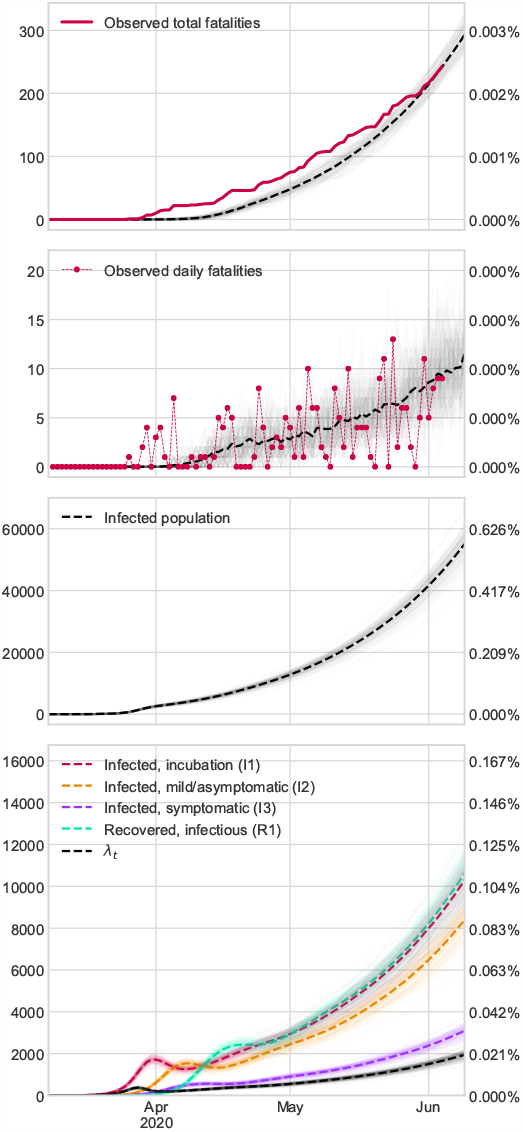
Results in Honduras: we assume that there are as many fatalities as in the official records.

**Figure 13:**
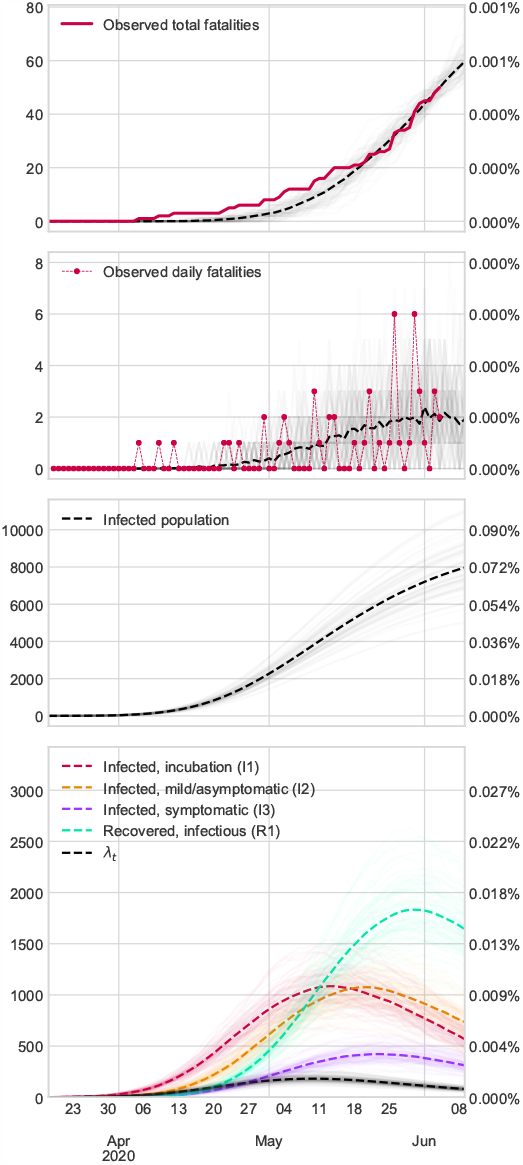
Results in Haití: we assume that there are as many fatalities as in the official records.

**Figure 14:**
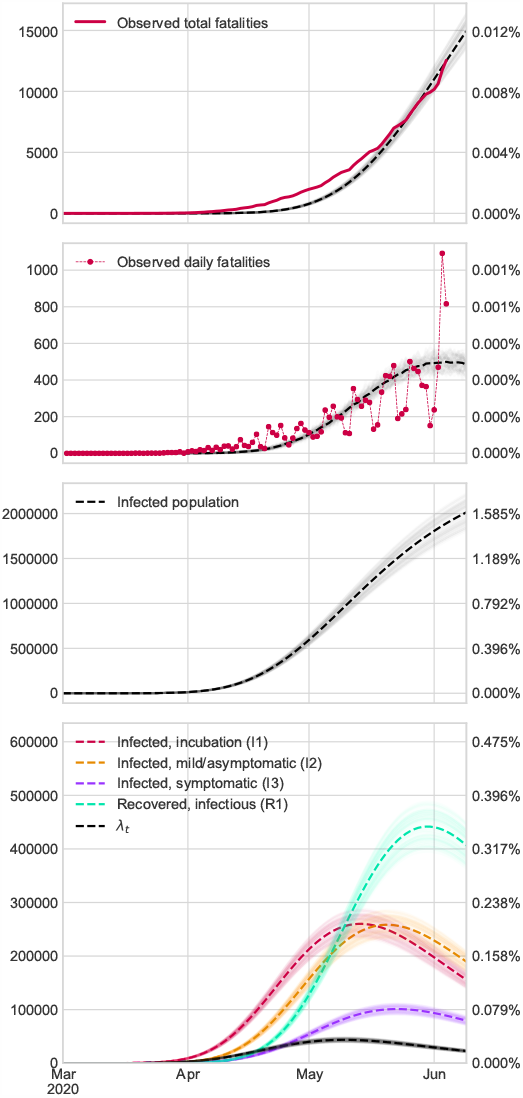
Results in Mexico: we assume that there are as many fatalities as in the official records.

**Figure 15:**
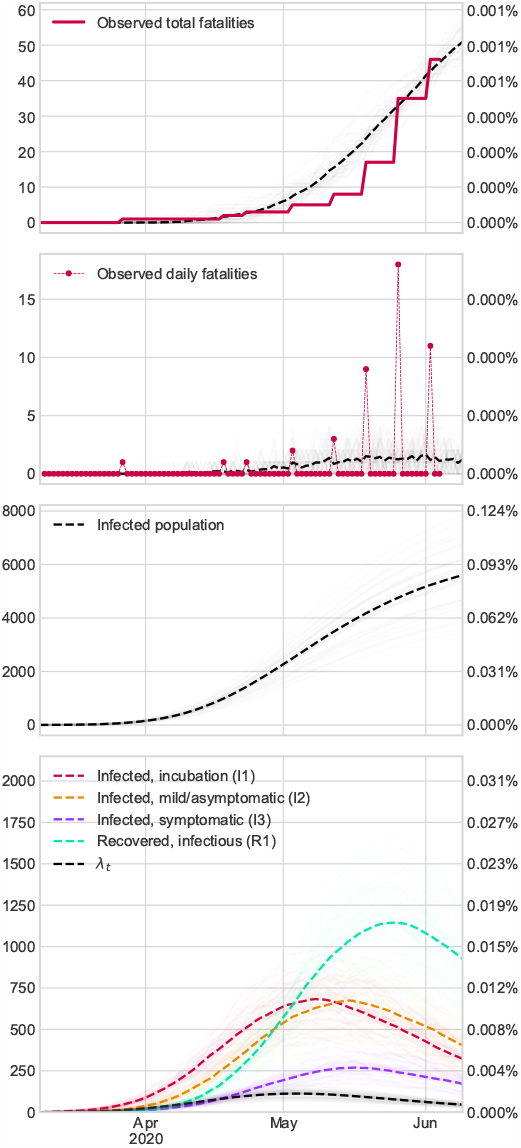
Results in Nicaragua: we assume that there are as many fatalities as in the official records.

**Figure 16:**
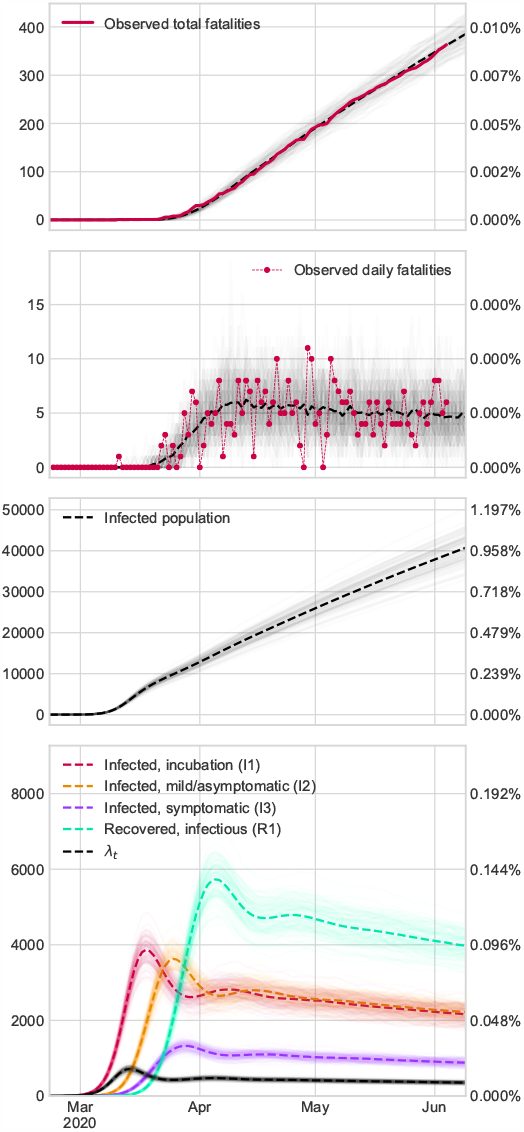
Results in Panamá: we assume that there are as many fatalities as in the official records.

**Figure 17:**
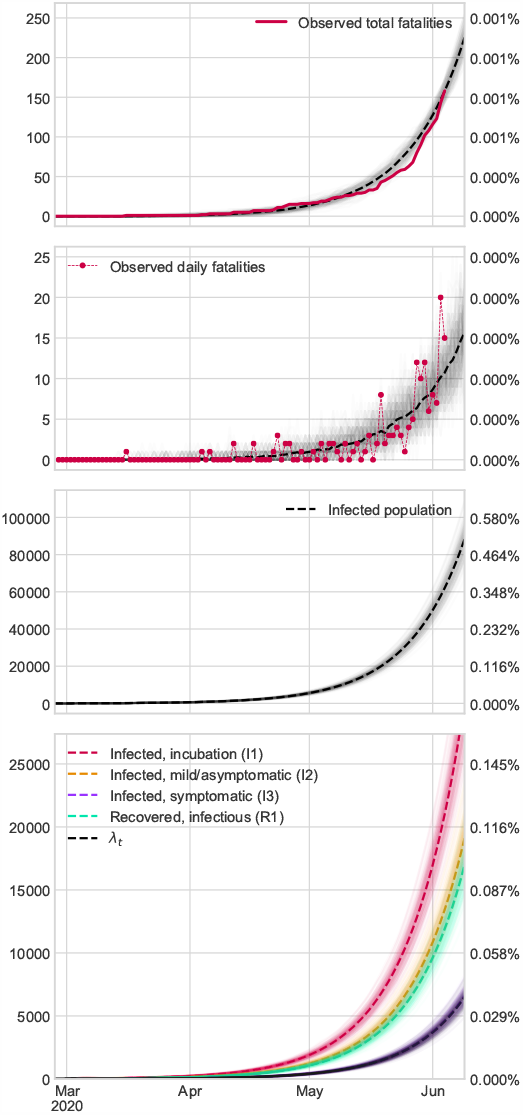
Results in Guatemala: we assume that there are as many fatalities as in the official records.

**Figure 18:**
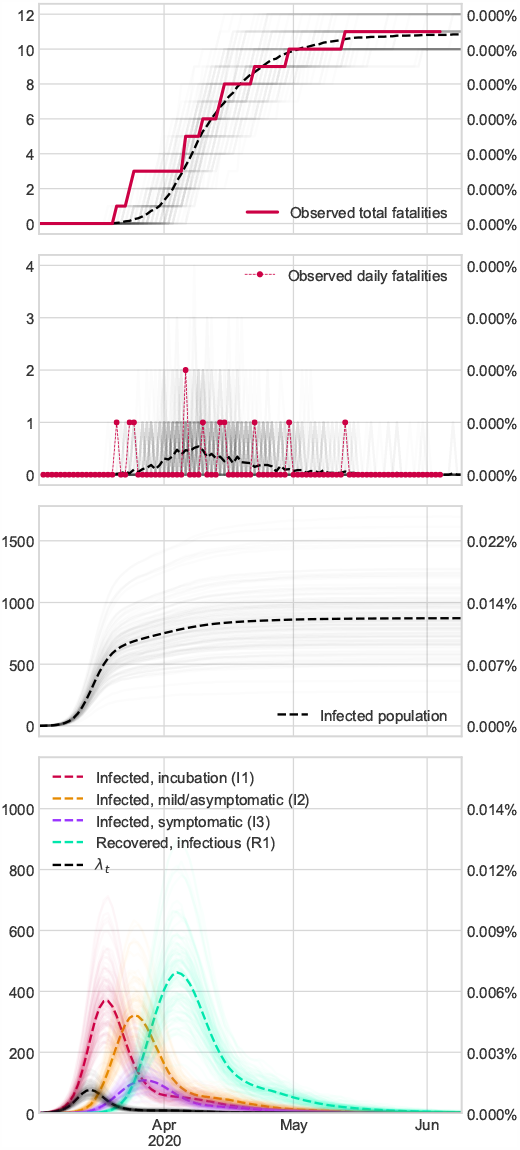
Results in Paraguay: we assume that there are as many fatalities as in the official records.

**Figure 19:**
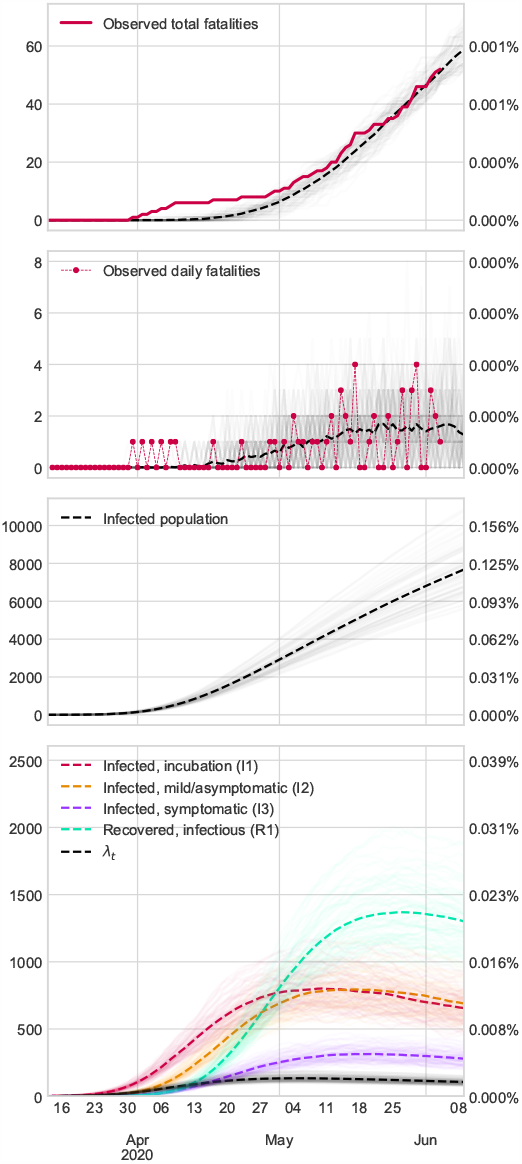
Results in El Salvador: we assume that there are as many fatalities as in the official records.

**Figure 20:**
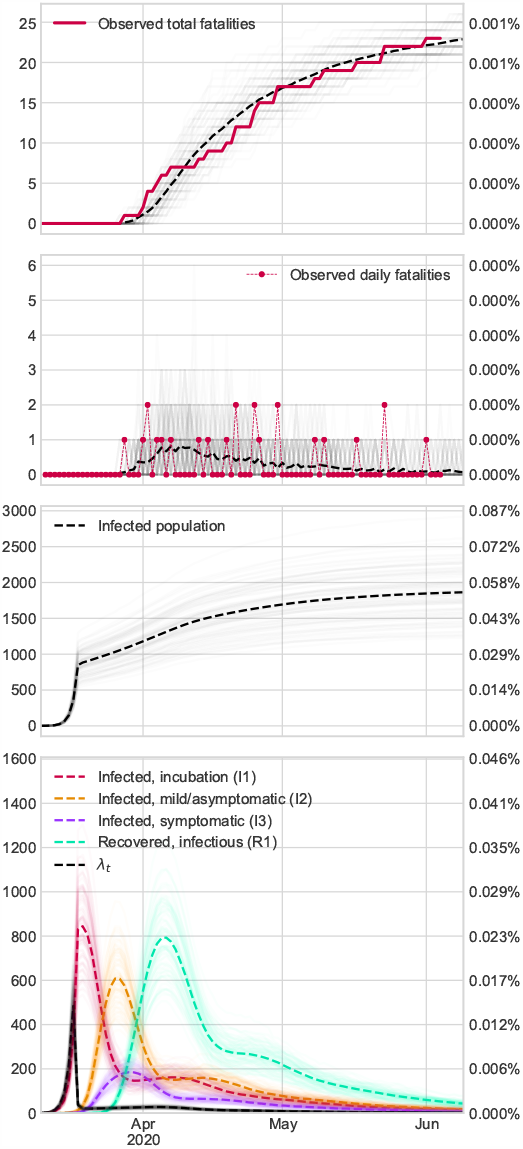
Results in Uruguay: we assume that there are as many fatalities as in the official records.

**Figure 21:**
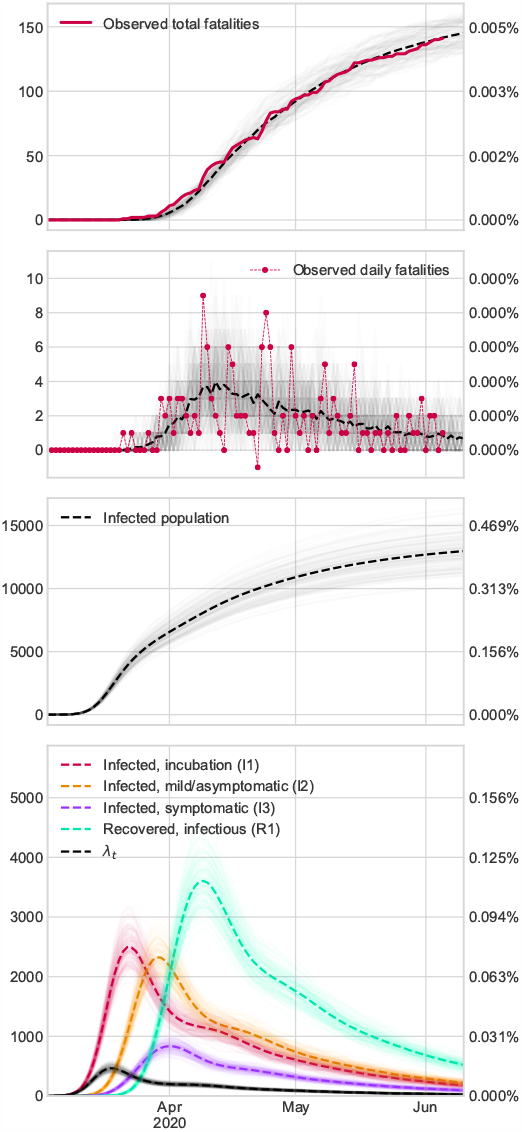
Results in Puerto Rico: we assume that there are as many fatalities as in the official records.

**Figure 22:**
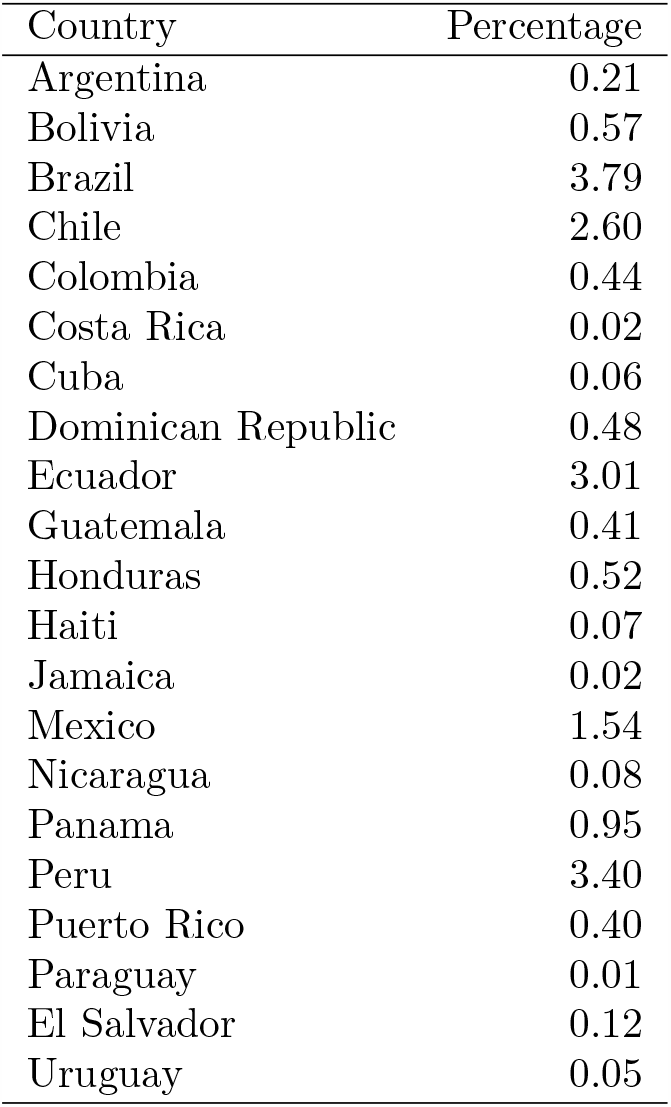
Estimated seroprevalence rate estimations in countries under study on June 6th

### 1.1 Methods

#### 1.1.1 Model parameters

Following the notation of our previosly pre-print [10], 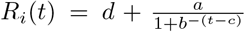, where *a ∈* [0, 5], *b ∈* [0, 1], *c ∈* [*−*30, 30], *d ∈* [0, 0.1]. The rest of parameter are tunning the same as original reference.

#### 1.1.2 Databases

Official data are obtained through John Hopkins repository (https://github.com/CSSEGISandData/COVID-19/tree/master/csse_covid_19_data/csse_covid_19_time_series).

## Supplementary Information

## Acknowledgements

This work has received financial support from Carlos III Health Institute, Grant/Award Number: COV20/00404, Ministry of Economy and Competitiveness (SPAIN) and the European Regional Development Fund (FEDER); and the Axencia Galega de Innovación, Consellería de Economía, Emprego e Industria, Xunta de Galicia, Spain, Grant/Award Number: GPC IN607B 2018/01. In addition, some member of this project has been partially support to develop this research by the Conseller??a de Cultura, Educación e Ordenación Universitaria (accreditation 2019-2022 ED431G-2019/04).

## Code availability

The code is publicly available from https://github.com/covid19-modeling/pyncov-19.

